# Emergency Department average wait time before admission, average time till sent home, and average number of violations for different hospitals types in Tennessee: Date Analysis of Medicare Data

**DOI:** 10.1101/2022.12.12.22283361

**Authors:** Viraj Brahmbhatt

## Abstract

The Emergency Department is a key facet of modern healthcare facilities and is responsible for bringing in a lot of patient volume. This healthcare asset is prone to a lot of shortcomings, primarily long waiting times. Though, patients primarily go to the emergency department in emergency scenarios, they still have a choice on which hospital they choose to attend. As such, it may be important for patients to be informed on the expected wait times at hospitals. Though the wait times vary each day, it may be important to know the average expected wait times, time till sent home, and number of violations by hospital type. Using the hospital data published by Medicare, the hospitals were grouped into 3 categories (Not for profit, For profit, and Academic) and data analysis was conducted and Analysis of Variance (ANOVA)was used to determine if the results were statistically significant. The analysis revealed that Academic hospital emergency departments had the highest time before admission, highest time until admission, and highest number of violations. The ANOVA revealed that the difference observed was significant in both time until admission (p-value=.0867) and number of violations (p-value = .011). The results of the study suggest that patients should consider attending a for-profit hospital emergency department if waiting time is a major concern. Analysis of the number of violations (indicative of poor care quality) suggested that for-profit hospitals do not have worse care quality and patients should not be worried about compromised quality care for the shorter wait time.

## Introduction

In the United States, there are variety of hospital system types that may be distinguished based on their ownership/operation type. These systems each have unique structural and managerial aspects that differentiate them. The primary hospital types are generally seen as Government owned, private nonprofit, private for profit, and academic. The quality of care received in these institutions may differ based on the key structural aspects that define the hospital type. For example, academic hospitals are known for having various tiers of providers and as such the thorough providence of care may come at the cost of a longer hospital visit.

The Emergency Department is a key facet of medical care and is marked as a positive contributor to the patient in-flow to a hospital system. The presence of an emergency room in a community may be seen as an indicator of better health outcomes. A 2007 publication by Nicholl et al. discussed distance to hospital and patient outcomes in emergency situations. The study found that patients living closer to a hospital tend to have better medical outcomes. For many, the emergency department serves as their primary source of medical care [1]. Though this is not advised, it further emphasizes the importance of the emergency department.

Patient’s choosing which emergency room they go to may affect their hospital experience. Just as hospital structure differs based on hospital type, the emergency department structure is also prone to difference. The effect of stress on decision making has been a long-studied topic. The literature reports that stress strongly affects decision making and causes an individual to not fully consider all the consequences of making a decision [2]. For patients in need of an emergency department, the stress brought on by sickness/injury may cause them to choose an emergency department without fully considering all aspects. A 2013 publication by Newgard et al. suggested that the decision of choosing an emergency department was heavily influenced by the patient and their families, even when transported by ambulance [3]. It is not known whether a specific hospital type would be better for emergency patient care than another.

There is literature present on how different hospital types correlate to differing patient outcomes. A 2002 publication by Devereaux et al. looking into mortality rates in private hospitals found that private for-profit hospitals possessed increased risk of death based on analysis of comorbidity and outlier adjusted mortality rates. Though mortality rates are not an indicator of quality care, it offers a hint that hospital type may in fact affect the type of care received. When it comes to choosing a hospital for care, academic hospitals are generally looked upon as being superior healthcare facilities. Typically, academic healthcare facilities will offer a wide range super specialized services and as a result patients associate them with better medical service [4]. There may be the concern with efficiency given the multi-tiered healthcare structure in medical facilities. A 2018 Kruse et al. looked into efficiency of hospitals by type in the European Union. The study found that public hospitals were not less efficient than private hospitals [5]. A study done on Medicare patients and the cost and quality of care depending on hospital type found that for-profit hospitals were generally associated with higher cost of care but there was not significant difference in outcome by hospital type [6].

The current emergency department structure has a major drawback in the form of long wait times and patient crowding. As reported by a 2015 publication by George and Evridiki, overcrowding of the emergency room is correlated with adverse patient outcomes as well as poor performances by providers [7]. In fact, the literature seems to indicate that patients view the overcrowding and resultant long wait times as negatives. A 2021 publication by Calder-Sprackman et al. suggested that patients like being able to know emergency department wait times prior to going to the hospital. In fact, patients felt greater quality care when they knew the waiting time [8]. Hospitals have taken note of the importance of wait times and are taking steps to reduce waiting times. One popular strategy being employed is a fast tracked approach to reduce wait times and to quickly address patients as they come in [9]. Strategies such as these are being tested to try and innovate the emergency department and improve patient experience in a medical facility.

## Methods

The data for the Emergency department wait times and violations in the past 10 years comes from the Medicare website which has publicly published the data. This data was sorted and processed through Microsoft Excel. The data for the hospitals ownership type is publicly published by each hospital system. Typically, there many categories of ownership that may be established (Government local, Government national, private Not for profit, Academic, Private For profit). For simplicity of categorization, three broad categories of Not for Profit, For Profit, and Academic were established and the hospitals were sorted into these 3 categories. These categories were established based on the operation structure rather than the ownership itself. This is because the study aimed to look into how the organizational structure affects wait time, time to discharge, and number of violations.

Once the data collection of hospital (emergency department) location, city population, wait time, time to discharge, number of violations in the past 10 years, and ownership type, the data was organized and the average value for wait time, time to discharge, number of violations in the past 10 years was found for each hospital type. Following this the values for wait time, time to discharge, number of violations in the past 10 years were plotted against city population for each of the hospitals. A linear regression was then conducted on this data to determine the strength of relationship. This was done to determine whether the population was a major contributor to longer wait times, longer time until discharge, and a greater number of violations. The results of the study may be useful in guiding consumer decision making.

## Results

Analysis of the average time before admission in the different hospital types revealed that Academic hospitals had the greatest wait time, while for profit hospitals had the shortest time before admission. Analysis of the average time till sent home revealed that academic hospitals had the greatest time, while for-profit hospitals had the lowest average time. Analysis of the average number of violations revealed that academic hospitals had the great number of violations, while not for profit hospitals had the lowest number of violations. Following this, a One-way Analysis of variance (Anova) was conducted to determine whether the difference observed among the groups was statistically significant. The Anova returned a p-value of .0867 for time until admission, .279 for time before admission, and .011 for number of violations. As such, academic hospitals possess significantly greater time before admission per an alpha level of .1. Additionally, academic hospitals possess a significantly greater number of violations as well. Analysis of the three measurements against population of the city did not reveal any significantly strong correlation. The plots of Number of violations, time till sent home, and time before admission were analyzed through linear regression. This technique helps to determine the strength of the relationship with the two variables The R^2^ value was .1154 for the number of violations against population. The R^2^ value was .2234 for time till sent home plotted against population. The R^2^ value was .0991 for time before admission

## Discussion

The results of the study suggest that there may be differences in the waiting time for the emergency department depending on the hospital attended. There is literature present on the correlations between longer wait times and lower perceived care quality in both the outpatient clinical setting and the emergency department [10]. Given this information, as well as the patient preference for knowing emergency department waiting times, the implications of this study may be useful in guiding consumer decision making [8]. Given that academic hospitals have displayed statistically greater wait times before admission as well as time till sent home, patients may choose to factor this into decision making prior to going into a hospital. The results from the 2013 Newgard et al. publication suggest that even in emergency situations patients still consider where they would like to be taken [3]. As such, the implications from this paper suggest that patients should consider going to a non for profit or a for profit hospital over an academic hospital.

There may be a variety of reasons beyond hospital structure that may have caused the wait times seen. Certain hospitals may see more complex patients and as a result the waiting times are greater. Additionally, For-profit hospitals were looked at negatively in patient-surveys and as such there may be lower patient volume at these facilities. This may have been responsible for the lower waiting time observed. The data from the Medicare hospital assessment website did not provide data on patient volume or patient difficulty. Future studies may choose to look into this as well as the average cost of being a patient at these hospitals as this may also serve as a deterrent to certain hospitals. There are many factors that may impact where a patient chooses to go. This study provided input on the wait times relative to another hospital type and what the patient may be able to expect.

## Data Availability

All data produced are available online at
https://www.medicare.gov/care-compare/?providerType=Hospital&redirect=true
and at each hospitals webpage.

